# Trajectories of child emotional and behavioural difficulties before and during the COVID-19 pandemic in a longitudinal UK cohort

**DOI:** 10.1101/2021.05.11.21257040

**Authors:** Elise Paul, Daphne Kounali, Alex S. F. Kwong, Daniel Smith, Ilaria Costantini, Deborah A. Lawlor, Kapil Sayal, Helen Bould, Nicholas J. Timpson, Kate Northstone, Melanie Lewcock, Kate Tilling, Rebecca M. Pearson

## Abstract

**Importance:** COVID-19 public health mitigation measures are likely to have detrimental effects on emotional and behavioural problems in children. However, longitudinal studies with pre-pandemic data are scarce.

**Objective:** To explore trajectories of children’s emotional and behavioural difficulties during the COVID-19 pandemic.

**Design and setting:** Data were from children from the third generation of a birth cohort study; the Avon Longitudinal Study of Parents and Children - Generation 2 (ALSPAC-G2) in the southwest of England.

**Participants:** The study population comprised of 708 children (median age at COVID-19 data collection was 4.4 years, SD=2.9, IQR= [2.2 to 6.9]), whose parents provided previous pre-pandemic surveys and a survey between 26 May and 5 July 2020 that focused on information about the COVID-19 pandemic as restrictions from the first lockdown in the UK were eased.

**Exposures:** We employed multi-level mixed effects modelling with random intercepts and slopes to examine whether children’s trajectories of emotional and behavioural difficulties (a combined total difficulties score) during the pandemic differ from expected pre-pandemic trajectories.

**Main outcomes:** Children had up to seven measurements of emotional and behavioural difficulties from infancy to late childhood, using developmentally appropriate scales such as the Emotionality Activity Sociability Temperament Survey in infancy and Strengths and Difficulties Questionnaire in childhood.

**Results:** The observed normative pattern of children’s emotional and behavioural difficulties pre-pandemic, was characterised by an increase in scores during infancy peaking around the age of 2, and then declining throughout the rest of childhood. Pre-pandemic, the decline in difficulties scores after age 2 was 0.6 points per month; but was approximately one third of that in post-pandemic trajectories (there was a difference in mean rate of decline after age 2 of 0.2 points per month in pre vs during pandemic trajectories [95 % CI: 0.10 to 0.30, *p* <0.001]). This lower decline in scores over the years translated to older children having pandemic difficulty scores higher than would be expected from pre-pandemic trajectories (for example, an estimated 10.0 point (equivalent of 0.8 standard deviations) higher score (95% CI: 5.0 to 15.0) by age 8.5 years). Results remained similar although somewhat attenuated after adjusting for maternal anxiety and age.

**Conclusion and relevance:** The COVID-19 pandemic may be associated with greater persistence of emotional and behavioural difficulties after the age 2. Emotional difficulties in childhood predict later mental health problems. Further evidence and monitoring of emotional and behavioural difficulties are required to fully understand the potential role of the pandemic on young children.

**Key Findings:** *Question:* How has the COVID-19 pandemic influenced emotional difficulties in young children?

*Findings:* Using repeated longitudinal data from before and during the pandemic we provide evidence that emotional difficulty scores of primary school aged children are higher by an estimated 10.0 points (0.8 standard deviations) (95% CI: 5.0 to 15.0) by age 8.5 years than would be expected based on pre pandemic data.

*Meaning:* The level of difference in emotional difficulties found in the current study has been linked to increased likelihood of mental health problems in adolescence and adulthood. Therefore, this increase in difficulties needs careful monitoring and support.

## Introduction

Public health measures adopted during the coronavirus disease 2019 (COVID-19) pandemic such as school and nursery closures, lockdown, and social distancing could have had detrimental effects on population mental health.^1^ Whilst children appear to be less affected physically by the COVID-19 virus itself, there are concerns that their mental health may be specifically affected by the pandemic mitigation measures.^2^ The pandemic occurred against a backdrop of rising mental health problems in children and adolescents, although the extent to which reflects increasing awareness and diagnoses is unclear.^3^

When asked to compare their children’s emotional and behavioural problems before and during the COVID-19 pandemic, around one-third of Spanish parents reported increases in children’s (ages 3-12) hyperactivity and conduct problems, but not emotional problems (e.g., unhappy, depressed).^4^ More than two-thirds of parents in another survey reported their children (ages 3-18) had more difficulty concentrating (76.6%), and were more nervous (38%) or more anxious (28.4%).^5^ Two cross-sectional studies of children aged 6-18 in China found that one in four and one in five met criteria for anxiety and depression, respectively.^6 7^ Whilst these cross-sectional findings are valuable, longitudinal research is needed.

To date, only a handful longitudinal studies have reported on changes in children’s mental health before and during the pandemic. A cohort of 168 children (Mean _age_ = 8.7) found increases in child-reported depressive symptoms, but not parent-reported emotional problems, compared to 18 months before the first UK lockdown.^8^ In another UK study, there was an 11% increase in both girls’ and boys’ aged 5-16 years probable mental disorder during as compared to before the pandemic.^9^ Similarly, a longitudinal study of 248 adolescents in Australia (Mean _age_ =14) reported increases in anxiety and depression symptoms during compared to before the pandemic.^10^ One UK study of 2988 children, aged 4-16 years, reported increases in hyperactivity/inattention and conduct problems from the beginning of the pandemic to July 2020.^11^ However, research on younger children is lacking.

The aim of this study was to examine whether the COVID-19 pandemic was associated with age-related trajectories of young children’s emotional and behavioural problems compared with expected trends, based pre-pandemic data. We use data from a questionnaire designed specifically for the COVID-19 pandemic in young children, nested within a UK open population cohort study The cohort we used enabled us to derive trajectories from infancy to late childhood during the pandemic and compare them to the same trajectories over the same ages in the same population. This provides a more robust method for estimating how young children have been affected by the pandemic, which is crucial for developing appropriate public health response.

## Methods

### Study design and participants

The Avon Longitudinal Study of Parents and Children (ALSPAC) is an ongoing longitudinal population-based study that recruited pregnant women residing in the south-west of England with expected delivery dates between 1^st^ April 1991 and 31^st^ December 1992.^12, 13^ The cohort consists of the original 13,761 mothers and their partners (G0) and their 14,901 children (G1).^14^ In 2012, ALSPAC began recruiting and collecting data on the next generation, G2, the children of the G1 participants and grandchildren of the originally recruited G0 women.^15^ Details of the age distribution according to data availability are provided in S1A and S1B.G2 participants can join the study at any time (from early pregnancy onwards), through an open cohort^15^. Therefore, the G2 children have a range of birthdates^15^ and ages at any wave of data collection (including the pre-pandemic and COVID data collection used here). More details of the participants who completed the COVID-19 survey are found here (https://wellcomeopenresearch.org/articles/5-278/v2).

Data are collected from both parents (at least one of whom is a G1 participant) and their children. The study website contains details of all data available through a fully searchable data dictionary (http://www.bristol.ac.uk/alspac/researchers/our-data/). Ethical approval for the study was obtained from the ALSPAC Law and Ethics Committee and the Local Research Ethics Committees and written informed consent was provided.

ALSPAC rapidly deployed two online-only questionnaires in response to the COVID-19 pandemic; one during the initial lockdown phase (9 April-15 May 2020), and the second when lockdown restrictions started to ease (26 May-5 July 2020). Data were collected using REDCap (Research Electronic Data CAPture tools),^16^ a secure web application for building and managing online data collection exercises hosted at the University of Bristol. As part of the second COVID-19 questionnaire, G1 parents were asked to complete an assessment about each of their G2 children. More details on the development of the questionnaire can be found elsewhere.^17^

This study includes 1,407 observations from 708 children (belonging to 522 families) with up to seven measures (six pre-pandemic and one during COVID-19). The mean number of measurements was 2.21 (SD = 1.32) and to be included children had to have at least one measure before or during the pandemic. The majority (*N* = 434; 83%) of the responding parents for this study was the mother. Pre-pandemic were collected between May 2012 and December 2018^15^ and questions about the COVID-19 pandemic data were collected in June 2020. A flow chart outlining the sample selection is presented in Figure S1 and further details of measures, including the number at each age, are provided in Figures S2-S5, and Tables S1A and S1B.

### Measures of child emotional and behavioural difficulties

Parents who participated in the COVID-19 survey completed one of two assessments regarding their child’s feelings and behaviour since the first UK lockdown began (23 March 2020), depending on the child’s age. Parents of children younger than 36 months (henceforth “younger children”) completed the mood and distractibility subscales of the Carey Infant Temperament Questionnaire (ITQ),^18^ and parents of those aged 36 months or older (“older children”) completed the Revised Rutter Parent Scale for Preschool Children.^19^ Measures which were the same as, or as close as possible to measuring the same underlying constructs, as the two used during the pandemic were selected for use in this study. Further information on each measure, including psychometric data, can be found in the Supplemental Materials.

#### Age 6 months (pre-pandemic) and ages 0-36 months (COVID-19 pandemic)

Summed scores of 19 parent-completed items from the ‘Mood’ and ‘Distractibility’ subscales of the 99 item ITQ^18^ were included.

#### Age 24 months (pre-pandemic)

Parents were asked 19 questions about their child’s recent behaviour from the ‘Mood’ and ‘Distractibility’ subscales of the Carey Toddler Temperament Questionnaire (TTQ).^20^

#### Age 48 months (pre-pandemic) and ages 36 months and upwards (COVID-19 pandemic)

Child emotional and behavioural difficulties were assessed using the behavioural difficulties score, which is a sum of the emotional difficulties, conduct difficulties, and the hyperactivity score from the Revised Rutter Parent Scale for Preschool Children.^19^

#### Ages 36, 60- & 72-months pre-pandemic

Parents completed the Emotionality Activity Sociability (EAS) Temperament Survey for Children^21^ which is comprised of four subscales corresponding to traits described by Buss and Plomin.^21^ Items from these four scales were summed to form the total scale, with higher scores indicating more difficulties: emotionality, activity, shyness, and sociability.

#### Age 84 months pre-pandemic

Twenty-five items from the parent-completed Strengths & Difficulties Questionnaire (SDQ)^22^ were summed to form the total difficulties score. This score total score includes assessing different aspects of the child’s mental health such as hyperactivity, conduct problems, and emotional difficulties.

### Measure of parental anxiety preceding the COVID-19 pandemic

We adjusted for parental anxiety to reduce bias associated with parent reports of their children’s emotional difficulties. We adjusted for anxiety rather than depression because our work on adult mental health in the parents found anxiety, rather than depression, increased during the pandemic.^23^ The most recently assessed before COVID-19 (age 24) measure of parental anxiety symptoms assessed using the Generalised Anxiety disorder (GAD) symptoms using a computerised version of the Clinical Interview Schedule-Revised (CIS-R)^24^ was included. We transformed this variable to an SD scale where 1 unit (1 SD) represents 3 points.

### Statistical analysis

We fitted three models using piecewise linear three-level mixed effects with random intercept and slopes to estimate trajectories for the within-child repeated measurements of emotional and behavioural difficulties over time (age), whilst accounting for clustering within families.^25^ The equation for each of the three models can be found in the Supplement. In Model 1, our characterisation of child emotional and behavioural difficulties trajectories are summarized by four parameters: i) the average score at age 2 where child difficulty scores peak, ii) the rate of score change before, iii) after 2 years, and iv) a quadratic term to account for non-linearity. To examine whether parental anxiety symptoms measured before the COVID-19 pandemic explained any patterns in children’s emotional and behavioural difficulties trajectories, we re-estimated Model 1 but including pre-pandemic parental anxiety (Model 2)

Within both models, we also examined the effect modification of ‘pandemic status’ on the trajectory parameters by including this as a two-level grouping variable with two options referring to whether the measure was assessed before or during the pandemic. Variance according to child gender was considered in all models as a further grouping variable. Analyses were conducted using Stata 16.^26^

### Missing data

139, 31% of the 522 parents, were missing parental anxiety measurements. We used multilevel multiple imputation using other measures of parental depression and anxiety that were collected pre-pandemic. We used ‘jomo’ for multilevel joint modelling multiple imputation.^27, 28^ We pooled parameter estimates from three-level mixed effect analyses adjusted for parental anxiety scores across 50 imputed datasets using Rubin’s rules.^29^

The objective of this study was to estimate the “total burden” of the pandemic on children’s emotional and behavioural well-bine. As COVID-19 mitigation measure are a universal exposure, and thus does not vary according to socio-demographic variables, we limited adjustments to parental anxiety and maternal age (because it associated with missing data) only.

## Results

Demographic characteristics of study children and their parents are provided in Table 1. Figure S2 depicts the observed trajectories of children’s emotional and behavioural difficulties scores with age. We examined and compared follow-up between those with and without observation during the pandemic as well as observed key socio-demographics (maternal age, at child’s birth, maternal education, quartile of multiple deprivation index, number of children, child’s sex, maternal anxiety). Follow-up patterns are also presented in the supplementary Figure S2. The association between having missing data on the COVID-19 survey and key socio-demographics was explored using mixed effects Poisson regression, we found no evidence for associations of child’s sex, and maternal parity, multiple area deprivation index or maternal education with completion of the COVID-19 survey (Table S8 and S9).

**Table 1.**
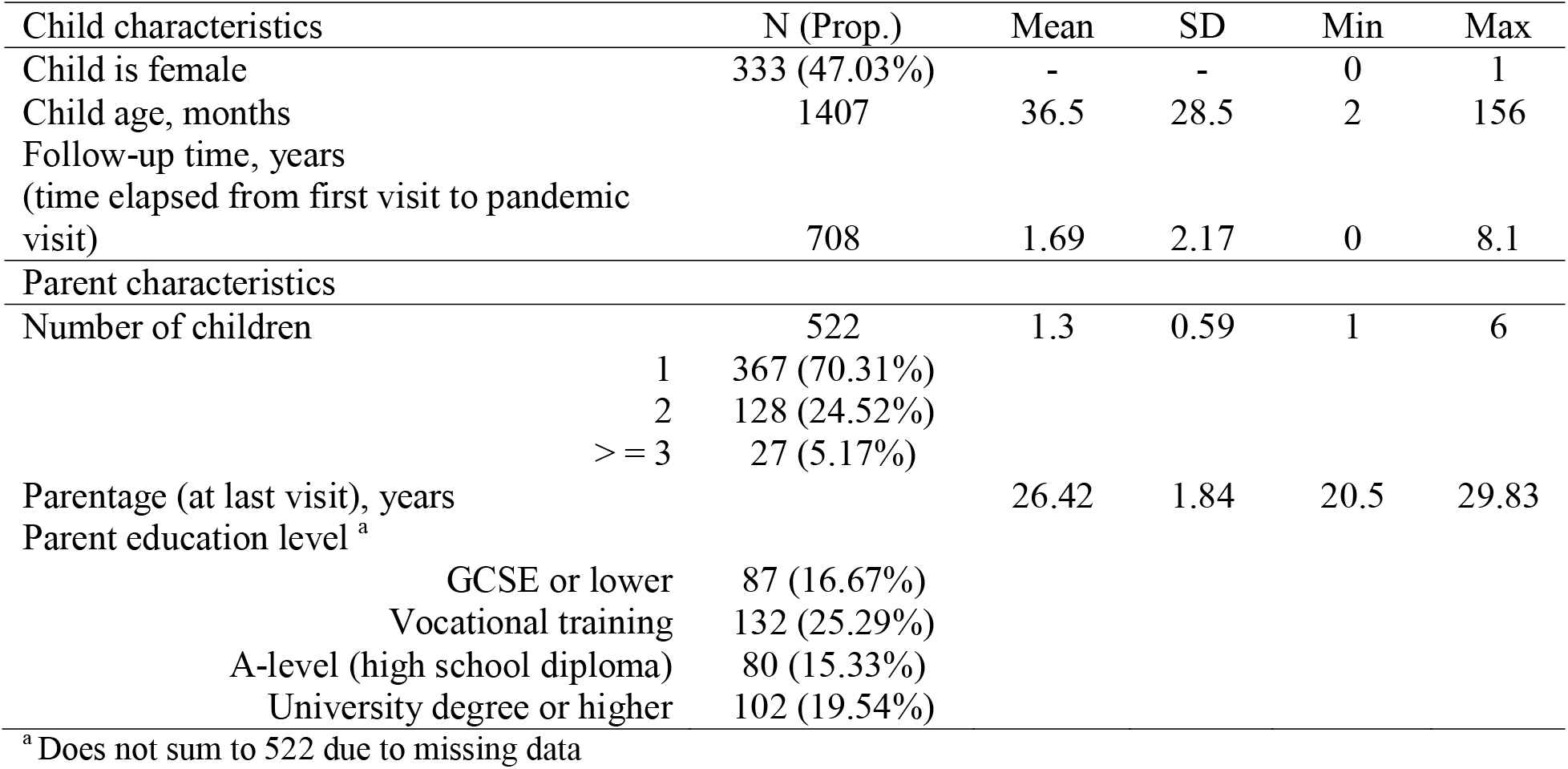
Demographic characteristics of children and their parents during the COVID-19 pandemic (*N* = 708)

Emotional and behavioural difficulties scores increase during infancy, peak around the age of 2 (24 months), and then decline during childhood at a decreasing rate of on average 0.565 (95% CI: 0.491 to 0.639) points per month (Table 2 and Figure 1). The peak difficulties scores at age 2 were lower during the COVID-19 pandemic compared with pre-pandemic scores for children at the same age. However, the rate of decline after age 2 was different according to pandemic status, where during-pandemic trajectories drop by approximately one-third as many points compared to pre-pandemic trajectories (average rate of decline= 0.195 (95% CI: 0.113 to 0.277)) (Figure 2 and Table S3). By age 8.5, there are differences of approximately 10 points (95% CI: 5.00 to 15.00) between pre- and during-pandemic scores, this represents a difference of 0.8 standard deviation.

**Figure 1.**
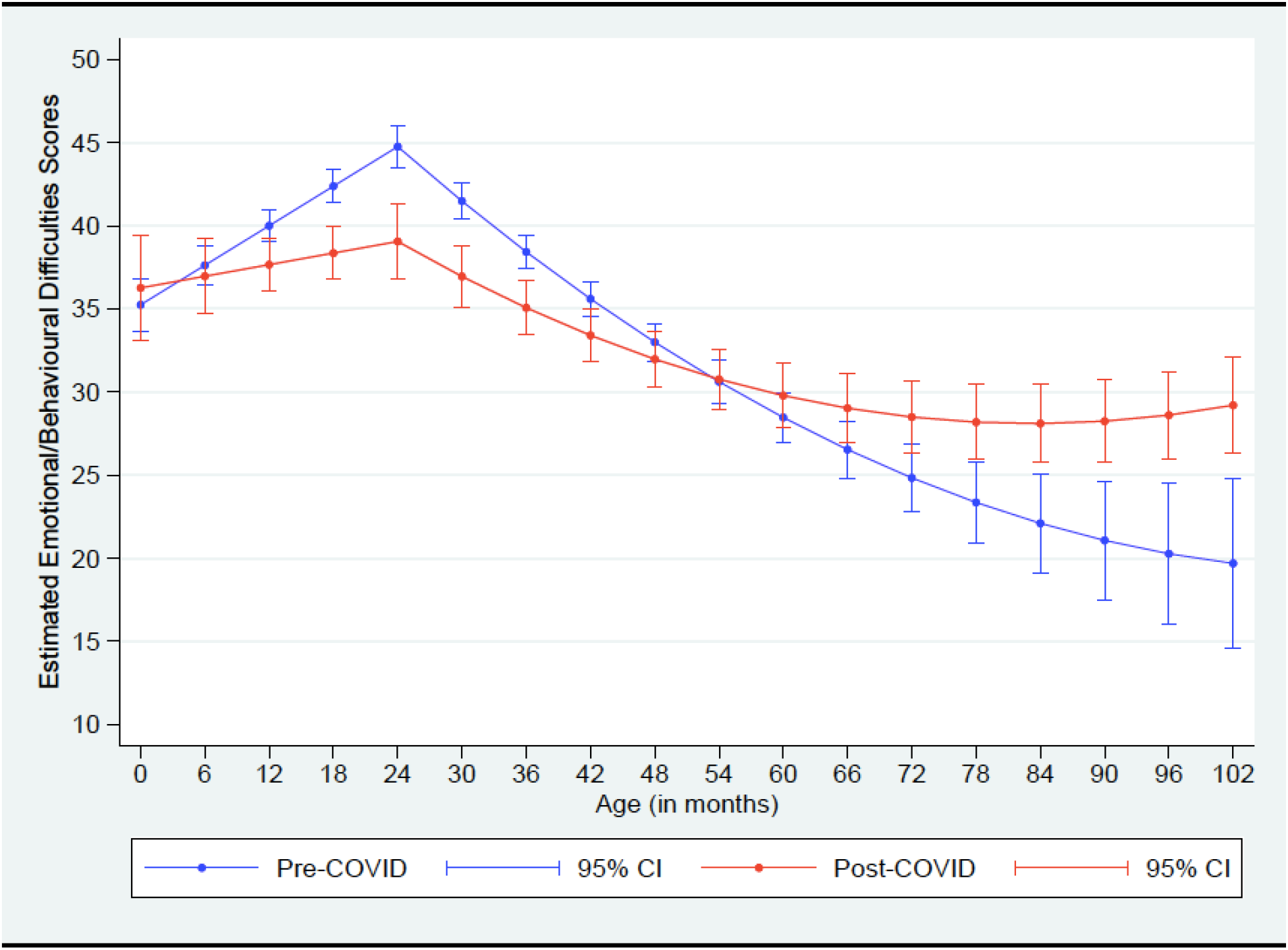
Predicted trajectories over time of children’s emotional and behavioural difficulties before and during (“post-COVID”) the COVID-19 pandemic.

**Figure 2.**
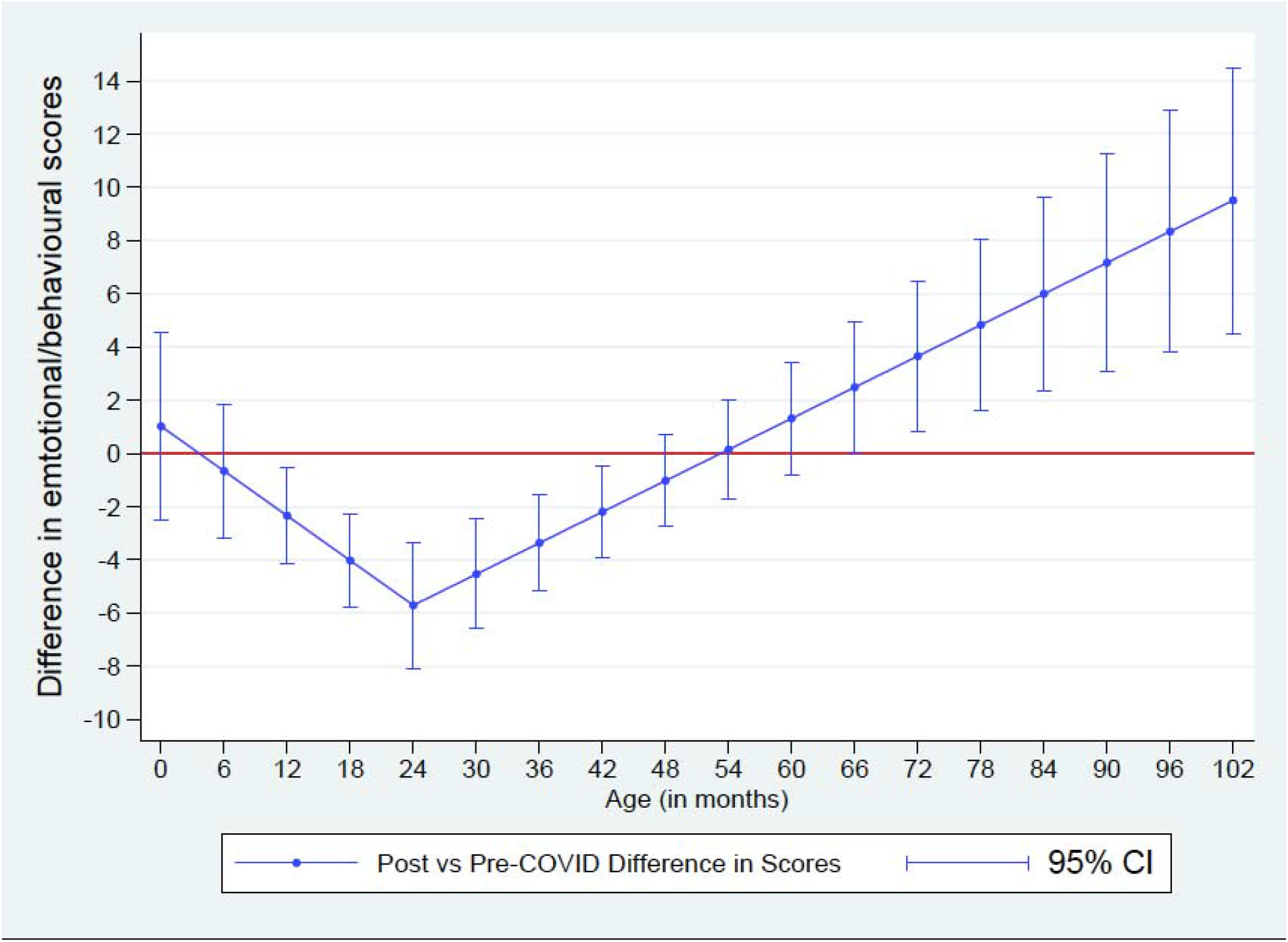
Estimated differences in total difficulty scores between during- minus pre-pandemic scores at each age.

**Table 2.**
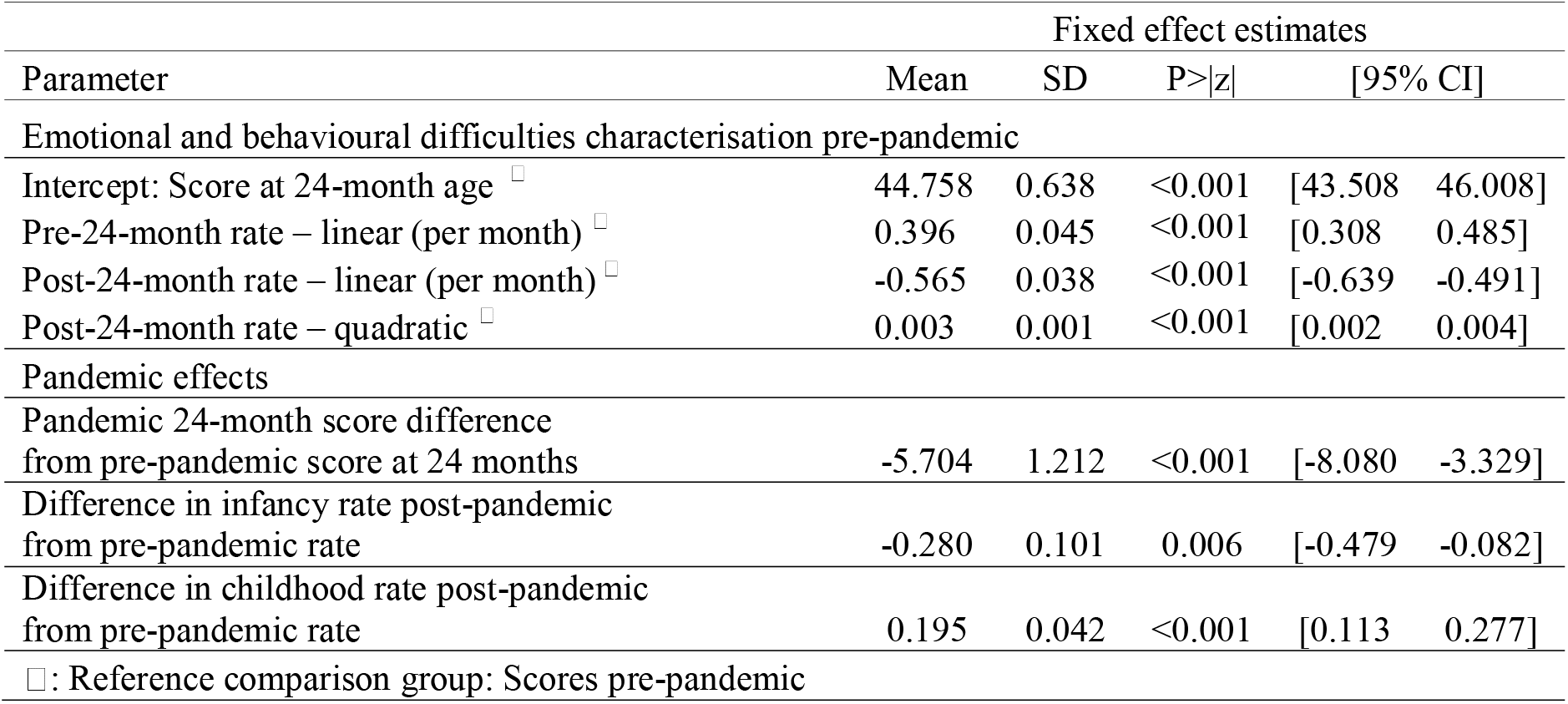
Model 1: Estimates from the three-level piecewise random effects (intercepts and slopes) model fitted to characterise emotional and behavioural difficulties score trajectories. (*N*= 708 children)

|  | Fixed effect estimates |  |  |  |  |
| --- | --- | --- | --- | --- | --- |
| Parameter | Mean | SD | P>\|z\| | [95% CI] |  |
| Emotional and behavioural difficulties characterisation pre-pandemic |  |  |  |  |  |
| Intercept: Score at 24-month age <sup>□</sup> | 44.758 | 0.638 | <0.001 | [43.508 | 46.008] |
| Pre-24-month rate – linear (per month) <sup>□</sup> | 0.396 | 0.045 | <0.001 | [0.308 | 0.485] |
| Post-24-month rate – linear (per month) <sup>□</sup> | -0.565 | 0.038 | <0.001 | [-0.639 | -0.491] |
| Post-24-month rate – quadratic <sup>□</sup> | 0.003 | 0.001 | <0.001 | [0.002 | 0.004] |
| Pandemic effects |  |  |  |  |  |
| Pandemic 24-month score difference from pre-pandemic score at 24 months | -5.704 | 1.212 | <0.001 | [-8.080 | -3.329] |
| Difference in infancy rate post-pandemic from pre-pandemic rate | -0.280 | 0.101 | 0.006 | [-0.479 | -0.082] |
| Difference in childhood rate post-pandemic from pre-pandemic rate | 0.195 | 0.042 | <0.001 | [0.113 | 0.277] |
| □: Reference comparison group: Scores pre-pandemic |  |  |  |  |  |

In Model 2, there was evidence for an association between parental anxiety and child trajectories (Table 3). For every 3-point increase (or 1 SD increase) in parental anxiety there was on average 1.060 increase in children’s difficulties scores (95% CI: 0.266 to 1.854). There was also evidence for a larger association between parental anxiety and trajectories during COVID-19 than before COVID-19 (Table S4). Adjustment for parental anxiety also attenuated differences in pre-pandemic and during pandemic trajectories but there was still evidence for a difference according to pandemic status where during pandemic trajectories declined by 0.073 points fewer than pre pandemic trajectories (95% CI: 0.004 to 0.142) (Table 3 and Figure S3). We adjusted for maternal age at child’s birth in both model 1 (Table S5) and model 2 including parental anxiety (Table S6), which had little influence on the results. In the model accounting for possible child gender interactions, the pandemic effect remains unchanged (Table S7). There was some weak evidence that boys may have higher scores in infancy but no evidence of gender differences in the rates of change in infancy or childhood. Results were similar for a model with the sample with no missing data (Table S10).

**Table 3.** Estimates from the three-level piecewise random effects (intercepts and slopes) model fitted to characterise emotional and behavioural difficulties score trajectories adjusted for pre-pandemic parental anxiety scores. (*N* = 708 children)

|  | Fixed effect estimates |  |  |  |  |
| --- | --- | --- | --- | --- | --- |
| Parameter | Mean | SD | P>\|z\| | [95% CI] |  |
| Emotional and behavioural difficulties characterisation pre-pandemic |  |  |  |  |  |
| Intercept: Score at 24-month age <sup>□</sup> | 44.412 | 0.644 | <0.001 | [43.150 | 45.673] |
| Pre-24-month rate – linear (per month) <sup>□</sup> | 0.375 | 0.046 | <0.001 | [0.284 | 0.466] |
| Post-24-month rate – linear (per month) <sup>□</sup> | -0.604 | 0.037 | <0.001 | [-0.677 | -0.530] |
| Post-24-month rate – quadratic <sup>□</sup> | 0.005 | 0.001 | <0.001 | [0.004 | 0.006] |
| Pandemic effects |  |  |  |  |  |
| Pandemic 24-month score difference from pre-pandemic score at 24 months | -3.476 | 1.166 | 0.003 | [-5.762 | -1.191] |
| Difference in infancy rate post-pandemic from pre-pandemic rate | -0.139 | 0.095 | 0.143 | [-0.324 | 0.047] |
| Difference in childhood rate post-pandemic from pre-pandemic rate | 0.073 | 0.035 | 0.037 | [0.004 | 0.142] |
| Parental anxiety scores |  |  |  |  |  |
| Parental anxiety (SD scale: 1 SD= 3 units) | 1.060 | 0.405 | 0.009 | [0.266 | 1.854] |
| □: Reference comparison group: Scores pre-pandemic |  |  |  |  |  |

## Discussion

In this third generation of a unique longitudinal birth cohort study in the UK, we found that children’s predicted trajectories of emotional and behavioural difficulties peaked around the age of two years, and then declined over the rest of early childhood. Emotional and behavioural difficulties in children under the age of two appeared to be lower during the pandemic than would be expected from pre-pandemic trajectories. However, the expected decline after age two in emotional and behavioural difficulties was considerably attenuated in during-pandemic trajectories compared to pre-pandemic trajectories, resulting in more difficulties at older ages during than before the pandemic. The projected difference of 0.8 SD is large with clear clinical relevance given that children with psychological difficulties are substantially more likely to go on to have a mental health disorder in adulthood.^30^ This work contributes to a small but growing body of evidence that the pandemic may be associated with increased emotional problems in children. ^8–11^

We found that parental anxiety was associated with higher child trajectories of emotional and behavioural problems in general (i.e., irrespective of pandemic status), but accounting for parental anxiety did not fully attenuate the results. Consistent with these findings, a review of the association between the current pandemic on mental health reported that parents of young children are disproportionately likely to affected by anxiety and depression symptoms.^31^ This increased parental stress associated with the pandemic is in turn, also likely to negatively influence child mental health via parenting practices,^32^ as well as parental substance misuse and domestic violence.^1^ One small study in the US found that greater COVID-19 disruptions and high anxious and depressive symptoms in parents of children under the age of 18 were associated with an increase in potential for child abuse.^32^ A Spanish study conducted early in the pandemic found evidence that children’s conduct and emotional problems were influenced by parents’ perceived pandemic-related distress mediated via parenting.^4^ Beyond a role of parental anxiety, children are likely to be negatively affected by lengthy and repeated school and nursery closures, which may result in a loss of structure and routine, support from school staff, time with peers, adequate nutrition, and physical activity.

Our available sample size for comparing pre- to during-pandemic child trajectories at very young ages was small and as a result, our estimates were less certain, as reflected in the wide confidence intervals. However, we had the benefit of up to seven repeated measures totalling more than >1,000 observations which increased power. Nonetheless, a larger sample size would be needed to examine how different facets of children’s functioning such as sub domains including hyperactivity or mood disturbances, may be driving our findings and explore different items within the broader constructs.

A further limitation is that temperament scales were used at younger ages (0-3) and emotion difficulty scales at later ages (3+). Evidence shows these two constructs are strongly linked,^34, 35^ however, there is a debate regarding the extent to which temperament is the same construct or a precursor of emotional difficulty.^33, 34^ Importantly, we have used equivalent constructs at each age in both pre and during the pandemic surveys and demonstrate that correlations between measures were similar to correlations between the same measure across ages. Thus, while caution is needed regarding interpretation of age-related change (e.g. the decline from 2 years might in part be due to differences in the two scales) at both time points, differences between pre and during the pandemic scores should not be markedly influenced by differences in the scales.

Given that the ALSPAC-G2 children are offspring of an original cohort born in the same period of time from 1991-1992, the age of the child during the pandemic is intrinsically linked to the age of their mother at their birth. By definition, older children have mothers who were younger when they were born. For example, a mother of a child who turned eight-years-old in 2020 would have been 21-22 years old at their birth. In contrast, a mother of a child who turned eight-years old in a 2017 pre-pandemic assessment would have been 18-19 years old at their birth. Given that young maternal age is a known risk factor for higher offspring emotional and behavioural problems,^35^ this is likely to have meant that pre-pandemic trajectories, especially for older children, would have been expected to be higher than during pandemic trajectories. This means if anything, the difference seen in older children during the pandemic in our study may be an underestimate. When we included parental age at birth of index child in the model this did not change the results. Nonetheless, it is important to acknowledge that the younger maternal age range and predominately white European ethnicity of our participants may limit generalisability to other, more diverse populations. Replication of our findings is important, though we are aware that few studies have the detailed repeat measures available in our study.

There are several implications for our findings. Emotional and behavioural difficulties early in childhood are associated with psychiatric disorder in later childhood^36^ and late adolescence.^37^ Our findings demonstrate a potential greater persistence of emotional and behavioural difficulties from the age of two onwards during the pandemic with much larger than expected scores by older ages. If replicated, this could be an early indication of potential risk of increases in rates of childhood psychiatric disorder in the population. It is therefore critical that parents and children are supported during further restrictions or lockdowns. Continued longitudinal research as the COVID-19 pandemic and its economic and social consequences continue to unfold will be necessary to determine if distress in children is maintained, as well as how presentations may change over time.

## Data Availability

The study website contains details of all data available through a fully searchable data dictionary (http://www.bristol.ac.uk/alspac/researchers/our-data/).

## Acknowledgements

We are extremely grateful to all the families who took part in this study, the midwives for their help in recruiting them, and the whole ALSPAC team, which includes interviewers, computer and laboratory technicians, clerical workers, research scientists, volunteers, managers, receptionists and nurses.

## Data Access

DK and EP had full access to all the data in the study and take responsibility for the integrity of the data and the accuracy of the data analysis.

## Supplemental Materials

### Measures of child emotional and behavioural difficulties

#### Age 6 months (pre-pandemic) and ages 0-36 months (COVID-19 pandemic)

Summed scores of 19 parent-completed items from the ‘Mood’ and ‘Distractibility’ subscales from the Carey Infant Temperament Questionnaire (ITQ),^1^ 99 item were included. Parents rated items on a 5-point scale, adopted from the original 6-point scale. The summed score of all 19 items indicates more emotional and behavioural difficulties. Internal consistencies were acceptable at both ages (Cronbach’s α = 0·66 and 0·70).

#### Age 24 months (pre-pandemic)

The Carey Toddler Temperament Questionnaire (TTQ)^2^ is designed for children aged 1-3 years and assesses nine temperamental characteristics. The 19 items included in the present study comprised the ‘Mood’ and ‘Distractibility’ subscales. The Mood (0·87) and Distractibility (0·69) subscales have been shown to have good test–re-test reliability.^2^ Parents rated statements on a 0-4 scale: almost never, rarely, sometimes, often, and almost always. As above, all 19 items were summed (Cronbach’s α= 0·64).

#### Age 48 months (pre-pandemic) and ages 36 months and upwards (COVID-19 pandemic)

Twenty-seven items from the 42-item Revised Rutter Parent Scale for Preschool Children^3^ were included. The present study used the behavioural difficulties score, which is a sum of the emotional difficulties, conduct difficulties, and the hyperactivity score, plus 10 other items, for a total of 27 items. Possible scores range from 0-54 points, with higher scores reflecting more difficulties (Cronbach’s α = 0·86).

#### Ages 36, 60- & 72-months pre-pandemic

Parents completed the 42-item Emotionality Activity Sociability (EAS) Temperament Survey for Children^4^ at ages 60 and 72 months, and 20 items at age 36 months pre-pandemic. The EAS is comprised of four subscales corresponding to traits described by Buss and Plomin:^4^ emotionality (tendency to show distress), activity (preferred level of activity), shyness (tendency to be inhibited with unfamiliar people), and sociability (tendency to prefer the company of others). Items are rated between 0 (almost never) and 4 (almost always). The EAS is designed to be used with children aged between 1-9 years old. The EAS has showed good reliability and stability over time.^5^ Items were summed to form the total scale, with higher scores indicating more difficulties. Internal reliability for all three ages was good (Cronbach’s α = 0·74-0·89).

#### Age 84 months pre-pandemic

Parents completed the Strengths & Difficulties Questionnaire (SDQ)^6^ when children were 84 months old pre-pandemic. The SDQ consists of 25-items assessing different aspects of the child’s mental health such as hyperactivity, conduct problems, and emotional difficulties. Parents are given four response options, from not true to true. Items were summed to form the total difficulties score (Cronbach’s α = 0·88).

We re-scaled child emotional and behavioural difficulties scores at different ages to a common scale (0-100) by dividing the sum score by the maximum possible in each scale and then multiplying by 100. See Tables S1A and S1B for basic descriptive information on each of the scales in original and standardised units.

Correlations between scales became stronger as age increased being 0.7-0.8 between ages 6, 7 and 8, and 0.3-0.4 between ages 2, 3, 4. The correlation between measures at age 3 (when a temperament scale was used) and age 4 (when an emotional/ behavioural scale was used) was larger than between ages 2 and 3 when temperament scales were used at both ages. This suggests the correlations across scales were similar to within scales at different ages.

**Table S1A.** Summary statistics for children’s pre-pandemic emotional and behavioural difficulties scores according to age.

| Pre-pandemic |  |  |  |  |  |  |  |  |  |  |  |  |
| --- | --- | --- | --- | --- | --- | --- | --- | --- | --- | --- | --- | --- |
| Emotional and Behavioural Difficulties Score |  |  |  |  |  |  |  |  |  |  |  |  |
| Measurement Wave |  | Original scale units |  |  |  |  | Transformed scale (0-100) |  |  |  |  |  |
|  | Measure | Age interval (months) observed | Mean Age | SD Age | No Items | Scale | Scale Max | N | Mean | SD | Min | Max |
| 6 months | ITS | (0-18) | 5.49 | 0.99 | 19 | 4 | 76 | 290 | 37.71 | 11.09 | 6.79 | 64.47 |
| 24 months | TTQ | (18-28) | 23.73 | 1.56 | 19 | 4 | 76 | 205 | 47.12 | 9.87 | 12.47 | 73.68 |
| 36 months | EAS <sup>a</sup> | (28-35) | 31.63 | 2.18 | 20 | 4 | 80 | 40 | 36.93 | 8.97 | 15.00 | 55.26 |
| 48 months | RR | (35-53) | 41.33 | 5.86 | 27 | 2 | 54 | 273 | 33.50 | 12.53 | 1.85 | 65.00 |
| 60 months | EAS <sup>b</sup> | (53-60) | 57.77 | 1.86 | 42 | 4 | 168 | 48 | 27.16 | 8.98 | 11.90 | 46.30 |
| 72 months | EAS <sup>b</sup> | (60-70) | 61.65 | 2.50 | 42 | 4 | 168 | 52 | 30.48 | 11.47 | 7.55 | 55.36 |
| 84 months | SDQ | >70 | 75.55 | 5.53 | 25 | 3 | 75 | 106 | 28.11 | 12.40 | 6.67 | 68.00 |

Note. ITS = Carey Infant Temperament Scale.<sup>1</sup> TTQ= Carey Toddler Temperament Questionnaire.<sup>2</sup> EAS = Emotionality Activity Sociability Temperament Survey for Children.<sup>4</sup> RR = Revised Rutter Parent Scale for Preschool Children.<sup>3</sup> SDQ = Strengths and Difficulties Questionnaire.<sup>6</sup> <sup>a</sup> Score contains 20 items from the EAS. <sup>b</sup> Score contains 42 items from the EAS.

**Table S1B.** Summary statistics for children’s emotional and behavioural difficulties scores during the COVID-19 pandemic according to age.

| During the COVID-19 pandemic |  |  |  |  |  |  |  |  |  |  |  |  |
| --- | --- | --- | --- | --- | --- | --- | --- | --- | --- | --- | --- | --- |
| Measurement Wave |  | Emotional and Behavioural Difficulties Score |  |  |  |  |  |  |  |  |  |  |
|  |  | Original scale units |  |  |  |  |  |  | Transformed scale 0-100 |  |  |  |
|  | Measure | Age interval (months) observed | Mean Age | SD Age | No items | Scale | Scale Max | N | Mean | SD | Min | Max |
| 0-36 months | ITS | (0-18) | 8.93 | 5.31 | 19 | 4 | 76 | 112 | 37.25 | 9.95 | 12.60 | 57.89 |
| 0-36 months | ITS | (18-28) | 22.57 | 2.67 | 19 | 4 | 76 | 60 | 38.88 | 10.39 | 13.50 | 55.26 |
| 0-36 months | ITS | (28-35) | 31.00 | 2.02 | 19 | 4 | 76 | 23 | 34.53 | 12.43 | 14.47 | 56.09 |
| 36 + months | RR | (35-53) | 45.47 | 5.14 | 27 | 2 | 54 | 49 | 30.46 | 10.81 | 11.11 | 53.70 |
| 36 + months | RR | (53-60) | 56.73 | 1.86 | 27 | 2 | 54 | 22 | 30.14 | 13.02 | 9.26 | 55.56 |
| 36 + months | RR | (60-70) | 63.91 | 3.12 | 27 | 2 | 54 | 22 | 35.28 | 17.60 | 8.57 | 64.20 |
| 36 + months | RR | >70 | 99.17 | 21.91 | 27 | 2 | 54 | 105 | 32.40 | 15.18 | 5.56 | 75.93 |

Note. ITS = Carey Infant Temperament Scale<sup>1</sup>, RR = Revised Rutter Parent Scale for Preschool Children<sup>3</sup>

**Table S2A.** Distribution of observations according to pre-pandemic age when measurements were undertaken.

| **Pre-pandemic** | **Maximum number of observations** |  |  |  |  |  |  |
| --- | --- | --- | --- | --- | --- | --- | --- |
| **Age interval**<br>**(months)**<br>**observed** | **1** | **2** | **3** | **4** | **5** | **6** | **7** |
| (0-18) | 206 | 84 | 58 | 30 | 10 | 12 | <5 |
| (18-28) | 68 | 24 | 18 | 10 | 5 | <5 | <5 |
| (28-35) | 14 | <5 | <5 | <5 | <5 | <5 |  |
| (35-53) | 31 | 23 | 14 | 9 | <5 | <5 |  |
| (53-60) | 9 | <5 | <5 | <5 |  |  |  |
| (60-70) | 7 | 7 | <5 |  |  |  |  |
| >70 | 30 | 7 |  |  |  |  |  |

**Table S2B.** Distribution of observations during the pandemic according to current age.

| During the pandemic |  | Maximum number of observations |  |  |  |  |  |
| --- | --- | --- | --- | --- | --- | --- | --- |
| Age interval |  |  |  |  |  |  |  |
| (months) |  |  |  |  |  |  |  |
| observed | 1 | 2 | 3 | 4 | 5 | 6 | 7 |
| (0-18) | 206 |  |  |  |  |  |  |
| (18-28) | 68 | 41 |  |  |  |  |  |
| (28-35) | 14 | 17 | 5 |  |  |  |  |
| (35-53) | 31 | 46 | 43 | 8 |  |  |  |
| (53-60) | 9 | 7 | 7 | 10 |  |  |  |
| (60-70) | 7 | 7 | 9 | 10 | <5 |  |  |
| >70 | 30 | 33 | 31 | 27 | 17 | 19 | <5 |

### Model 1 Equation

The model assumes that score measurements depend upon the critical age of 24 months such that each subject has a baseline level and a pre- and post-critical age slope. If we denote *a_ijt_* the age at 24 months for each child j from responder/carer i, at each measurement occasion t, then

*Y_ijt_* is the jth child score from parent i at each measurement occasion t

*T_ijt_* = 24 – *a_ijt_* is the timing of the t^th^ measurement before and after the critical age.

*δ_ijt_* is an indicator variable taking the of value 1 of the individual child measurement occasion is before the age of 24 months and 0 otherwise.

*X_jt_* is an indicator variable taking the of value 1 of the individual child measurement occasion is taken before the pandemic and 0 otherwise.

*S_j_* is an indicator variable taking the of value 1 of the individual child is a girl and 0 if child is a boy.

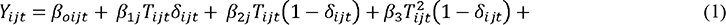

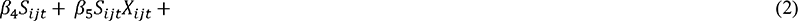

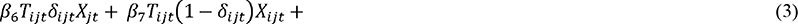

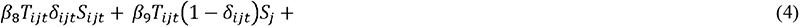

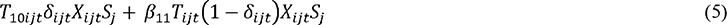

where,

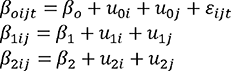

*ε_ijt_* is residual error (including measurement error normally distributed) and *u_kij_*, k=0,1,2 for carer i and individual child j, are the random effects terms, independently and normally distributed and correspond to the intercept and pre- and post-critical age slopes, respectively and partitioning the different variance components corresponding to the level of carer/mother and individual child.

Part (1) of the model equation above correspond to over trajectory characterised by an intercept term representing the score level at the critical age of 24 months and pre- and post-critical age along with an additional quadratic term post critical age. These terms represent boys’ pre-pandemic.

Part (2) These terms represent how the intercept is modified by the pandemic is boys and girls (interactions)

Part (3) These terms represent how the pre- and post-critical age slopes are modified by sex (interactions)

Part (4) These terms represent how the pre- and post-critical age slopes are modified by the pandemic in boys (interactions)

Part (5) These terms represent how the pre- and post-critical age slopes are modified by the pandemic in girls (interactions)

Adjustments for maternal anxiety can be readily incorporated in the above equation as an additional predictor.

**Table S3.** Unadjusted differences in post vs pre differences in difficulties scores taken from the model in Figure 1.

| **Age (years)** | **Mean**<br>**Difference** | **SD** | **Low CI** | **High CI** |
| --- | --- | --- | --- | --- |
| 0 | 1.02 | 1.79 | -2.49 | 4.53 |
| 0.5 | -0.66 | 1.29 | -3.19 | 1.86 |
| 1 | -2.34 | 0.93 | -4.16 | -0.53 |
| 1.5 | -4.02 | 0.89 | -5.77 | -2.28 |
| 2 | -5.70 | 1.21 | -8.08 | -3.33 |
| 2.5 | -4.53 | 1.05 | -6.59 | -2.47 |
| 3 | -3.36 | 0.93 | -5.19 | -1.54 |
| 3.5 | -2.19 | 0.87 | -3.89 | -0.50 |
| 4 | -1.02 | 0.87 | -2.73 | 0.69 |
| 4.5 | 0.15 | 0.95 | -1.71 | 2.00 |
| 5 | 1.32 | 1.08 | -0.79 | 3.43 |
| 5.5 | 2.49 | 1.24 | 0.05 | 4.92 |
| 6 | 3.66 | 1.43 | 0.85 | 6.47 |
| 6.5 | 4.83 | 1.64 | 1.61 | 8.05 |
| 7 | 6.00 | 1.86 | 2.36 | 9.64 |
| 7.5 | 7.17 | 2.08 | 3.09 | 11.26 |
| 8 | 8.34 | 2.31 | 3.80 | 12.88 |
| 8.5 | 9.51 | 2.55 | 4.52 | 14.51 |

**Table S4.** Estimates from the three-level piecewise random effects (intercepts and slopes) model fitted to characterise emotional and behavioural difficulties score trajectories Adjusted for pre-pandemic parental anxiety scores and interaction with COVID-19 (N = 708 children).

| Parameter | Fixed effect estimates |  |  |  |  |
| --- | --- | --- | --- | --- | --- |
|  | Mean | SD | P>\|z\| | [95% CI] |  |
| Emotional and behavioural difficulties characterisation pre-pandemic |  |  |  |  |  |
| Intercept: Score at 24-month age <sup>□</sup> | 44.446 | 0.644 | <0.001 | [43.184 | 45.708] |
| Pre-24-month rate – linear (per month) <sup>□</sup> | 0.375 | 0.046 | <0.001 | [0.284 | 0.466] |
| Post-24-month rate – linear (per month) <sup>□</sup> | -0.603 | 0.037 | <0.001 | [-0.676 | -0.530] |
| Post-24-month rate – quadratic <sup>□</sup> | 0.005 | 0.001 | <0.001 | [0.004 | 0.006] |
| Pandemic effects |  |  |  |  |  |
| Pandemic 24-month score difference from pre-pandemic score at 24 months | -3.514 | 1.169 | 0.003 | [-5.805 | -1.223] |
| Difference in infancy rate post-pandemic from pre-pandemic rate | -0.145 | 0.095 | 0.126 | [-0.331 | 0.041] |
| Difference in childhood rate post-pandemic from pre-pandemic rate | 0.072 | 0.035 | 0.040 | [0.003 | 0.141] |
| Parental anxiety scores (SD scale 1 SD = 3 units) |  |  |  |  |  |
| Parental anxiety effect pre-pandemic | 0.809 | 0.461 | 0.080 | [-0.096 | 1.714] |
| Parental anxiety effect post- pandemic | 1.716 | 0.695 | 0.014 | [0.352 | 3.081] |
| □: Reference comparison group: Scores pre-pandemic |  |  |  |  |  |

**Table S5.** Estimates from the three-level piecewise random effects (intercepts and slopes) model fitted to characterise emotional and behavioural difficulties score trajectories Adjusted for maternal age at child’s birth (N = 708 children)

|  | Fixed effect estimates |  |  |  |  |
| --- | --- | --- | --- | --- | --- |
| Parameter | Mean | SD | P>\|z\| | [95% CI] |  |
| Emotional and behavioural difficulties characterisation Pre-pandemic |  |  |  |  |  |
| Intercept: Score at 24-month age <sup>□</sup> | 44.602 | 0.659 | <0.001 | [43.309 | 45.894] |
| Pre-24-month rate – linear (per month) <sup>□</sup> | 0.390 | 0.046 | <0.001 | [0.301 | 0.479] |
| Post-24-month rate – linear (per month) <sup>□</sup> | -0.575 | 0.039 | <0.001 | [-0.651 | -0.499] |
| Post-24-month rate – quadratic <sup>□</sup> | 0.003 | 0.001 | <0.001 | [0.002 | 0.005] |
| Pandemic effects |  |  |  |  |  |
| Pandemic 24-month score difference from pre-pandemic score at 24 months | -5.284 | 1.288 | <0.001 | [-7.809 | -2.759] |
| Difference in infancy rate post-pandemic from pre-pandemic rate | -0.288 | 0.102 | 0.005 | [-0.487 | -0.089] |
| Difference in childhood rate post-pandemic from pre-pandemic rate | 0.189 | 0.042 | 0.000 | [0.107 | 0.272] |
| Maternal age at child’s birth (SD scale 1 SD = 3 units) | -0.471 | 0.502 | 0.348 | [-1.455 | 0.513] |
| □: Reference comparison group: Scores pre-pandemic |  |  |  |  |  |

**Table S6.**
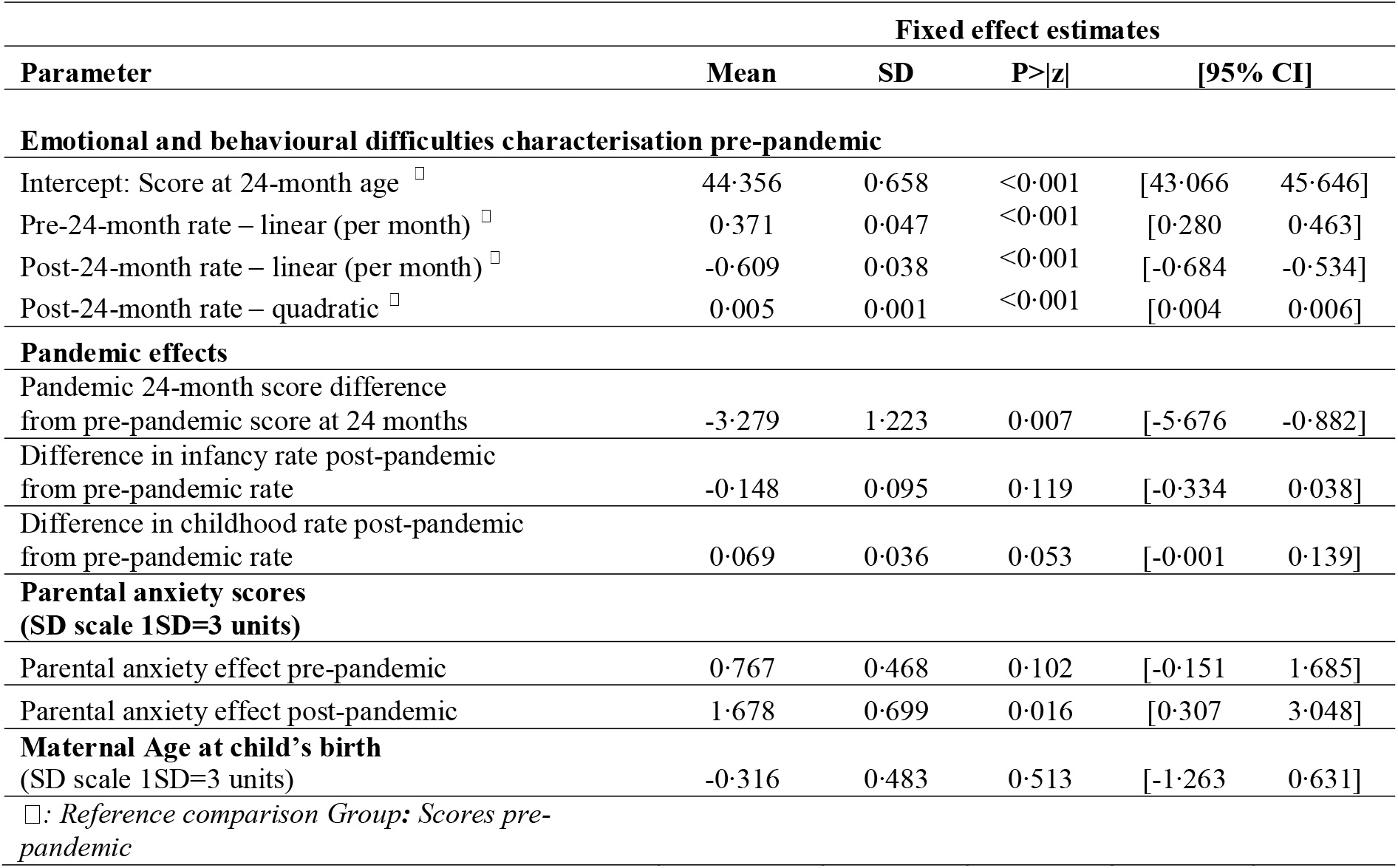
Estimates from the three-level piecewise random effects (intercepts and slopes) model fitted to characterise emotional and behavioural difficulties score trajectories Adjusted for pre-pandemic parental anxiety scores and interaction with COVID-19 and maternal age at child’s birth (N = 708 children contributing 1407 observations reported by 525 parents: after multiple imputation for maternal anxiety)

**Table S7.** Model 1: Estimates from the three-level piecewise random effects (intercepts and slopes) model fitted to characterise emotional and behavioural difficulties score trajectories. (N = 708 children) adjusted for potential sex differences in trajectories.

| Fixed effect estimates |  |  |  |  |  |
| --- | --- | --- | --- | --- | --- |
| Parameter | Mean | SD | P>\|z\| | [95% CI] |  |
| Emotional and behavioural difficulties characterisation Pre-pandemic |  |  |  |  |  |
| Intercept: Score at 24-month age <sup>□</sup> | 45.621 | 0.782 | <0.001 | [44.089 | 47.153] |
| Pre-24-month rate – linear (per month) <sup>□</sup> | 0.446 | 0.057 | <0.001 | [0.334 | 0.559] |
| Post-24-month rate – linear (per month) <sup>□</sup> | -0.557 | 0.041 | <0.001 | [-0.638 | -0.476] |
| Post-24-month rate – quadratic <sup>□</sup> | 0.003 | 0.001 | <0.001 | [0.002 | 0.004] |
| Pandemic effects |  |  |  |  |  |
| Pandemic 24-month score difference from pre-pandemic score at 24 months | -5.652 | 1.209 | <0.001 | [-8.021 | -3.283] |
| Difference in infancy rate post-pandemic from pre-pandemic rate | -0.280 | 0.101 | 0.006 | [-0.478 | -0.081] |
| Difference in childhood rate post-pandemic from pre-pandemic rate | 0.190 | 0.042 | <0.001 | [0.109 | 0.272] |
| Pre-pandemic gender differences |  |  |  |  |  |
| Girl 24-month score difference From boys at 24 months | -1.907 | 1.002 | 0.057 | [-3.872 | 0.057] |
| Girl infancy rate difference From boys at 24 months | -0.109 | 0.078 | 0.163 | [-0.262 | 0.044] |
| Girl childhood rate difference From boys at 24 months | -0.010 | 0.038 | 0.798 | [-0.084 | 0.064] |
| □: Reference comparison Group: Scores pre-pandemic |  |  |  |  |  |

**Table S8.**
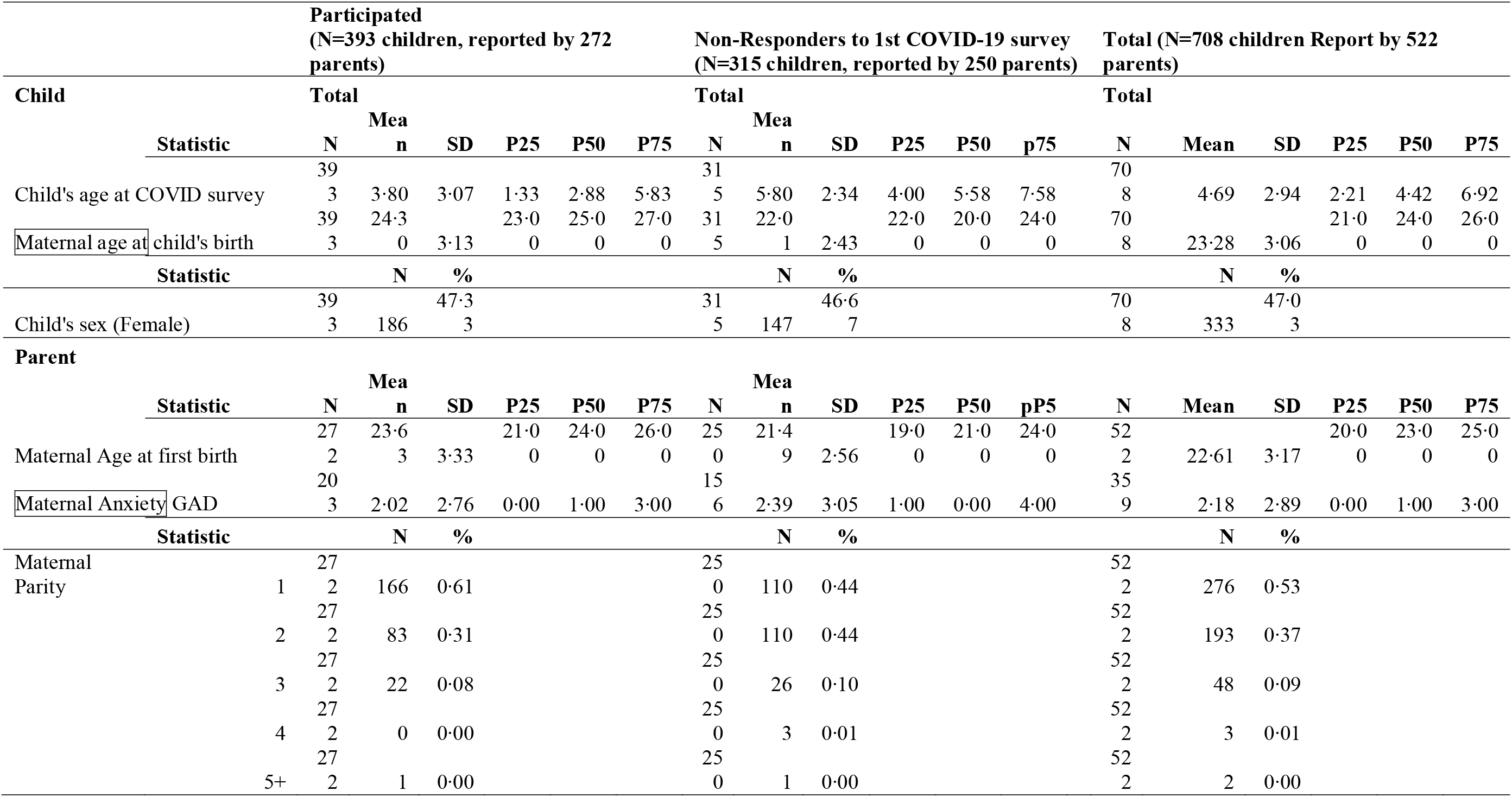
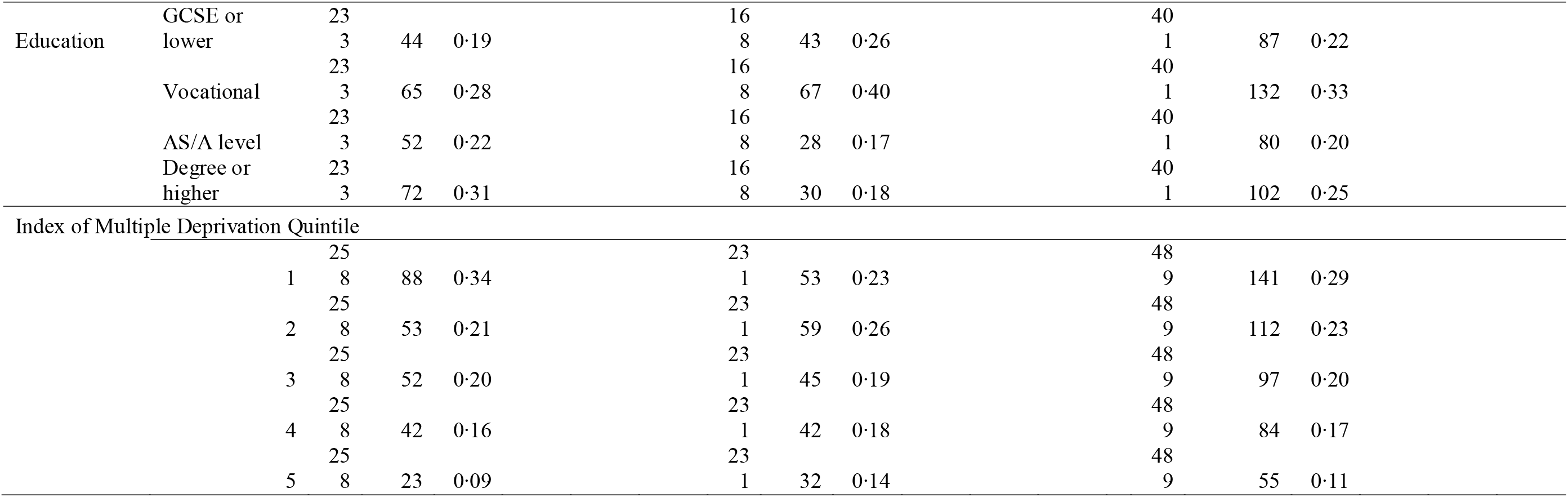
Description of Key Socio-demographics characteristics according to response to the COVID survey.

**Table S9:** Mixed effects Poisson Regression examining the dependence between missing rates and key child and parental sociodemographic characteristics including response to COVID survey·.

| Characteristic | Units | Reference | IRR<sup>y</sup> | SE | P-value | 95% CI |  |
| --- | --- | --- | --- | --- | --- | --- | --- |
| **Child** |  |  |  |  |  |  |  |
| Child Sex | 0-1 | Male | 0.88 | 0.08 | 0.171 | [0.73 | 1.06] |
| **Parent** |  |  |  |  |  |  |  |
| COVID-Survey Non-Responder | 0-1 | Responded | 1.54 | 0.15 | <0.001 | [1.28 | 1.87] |
| Maternal age at first birth | years |  | 2.09 | 0.04 | <0.001 | [2.02 | 2.17] |
| Parent's sex | 0-1 | Male | 0.90 | 0.12 | 0.420 | [0.70 | 1.16] |
| Parity | 2 | 1 child | 1.20 | 0.12 | 0.075 | [0.98 | 1.47] |
|  | +3 |  | 1.04 | 0.17 | 0.786 | [0.76 | 1.43] |
| Maternal education | Vocational | GCSE or lower | 0.81 | 0.10 | 0.098 | [0.63 | 1.04] |
|  | AS/A level |  | 0.89 | 0.13 | 0.427 | [0.67 | 1.19] |
|  | Degree or higher |  | 0.99 | 0.14 | 0.935 | [0.74 | 1.31] |
| Index of Multiple Deprivation Quantile | 2 | First Quantile | 0.86 | 0.12 | 0.281 | [0.66 | 1.13] |
|  | 3 |  | 0.84 | 0.12 | 0.201 | [0.64 | 1.10] |
|  | 4 |  | 1.09 | 0.16 | 0.573 | [0.81 | 1.45] |
|  | 5 |  | 0.91 | 0.16 | 0.577 | [0.65 | 1.27] |
| Maternal Anxiety | 0-12 |  | 1.02 | 0.02 | 0.251 | [0.99 | 1.05] |

<sup>y</sup>IRR: incidence rate ratio
<sup>‡</sup>IRR: incidence rate ratio
Missing rates are higher among non-responders in the COVID survey where non-responders have 1.5 times the missing rate of responders. Apart from maternal age we did not find any strong evidence of association with other parental key demographics.

**Table S10.**
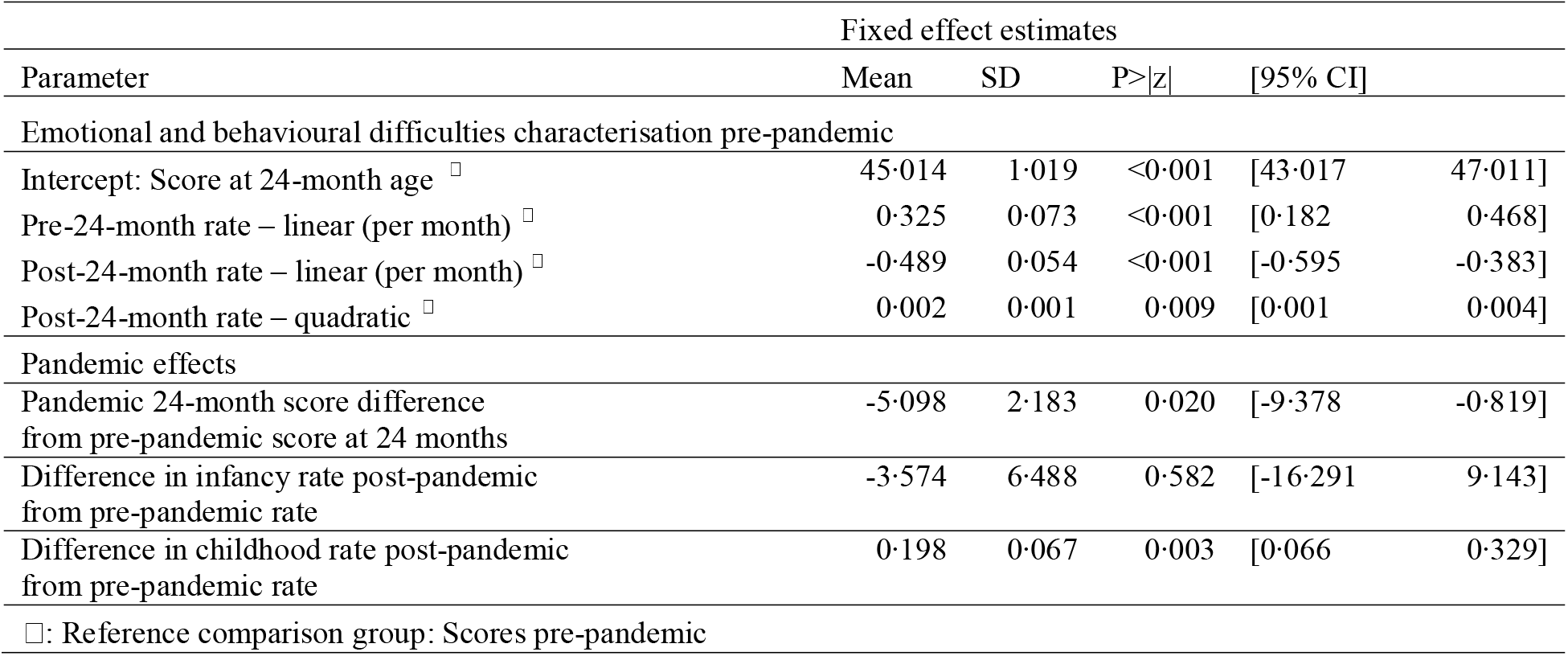
Model 1: Estimates from the three-level piecewise random effects (intercepts and slopes) model fitted to characterise emotional and behavioural difficulties score trajectories for those children with were observed during the pandemic and also had pre-pandemic observations. (N= 188 children i.e., excluding those with no pre-covid or no-post-covid observation)

**Figure S1.**
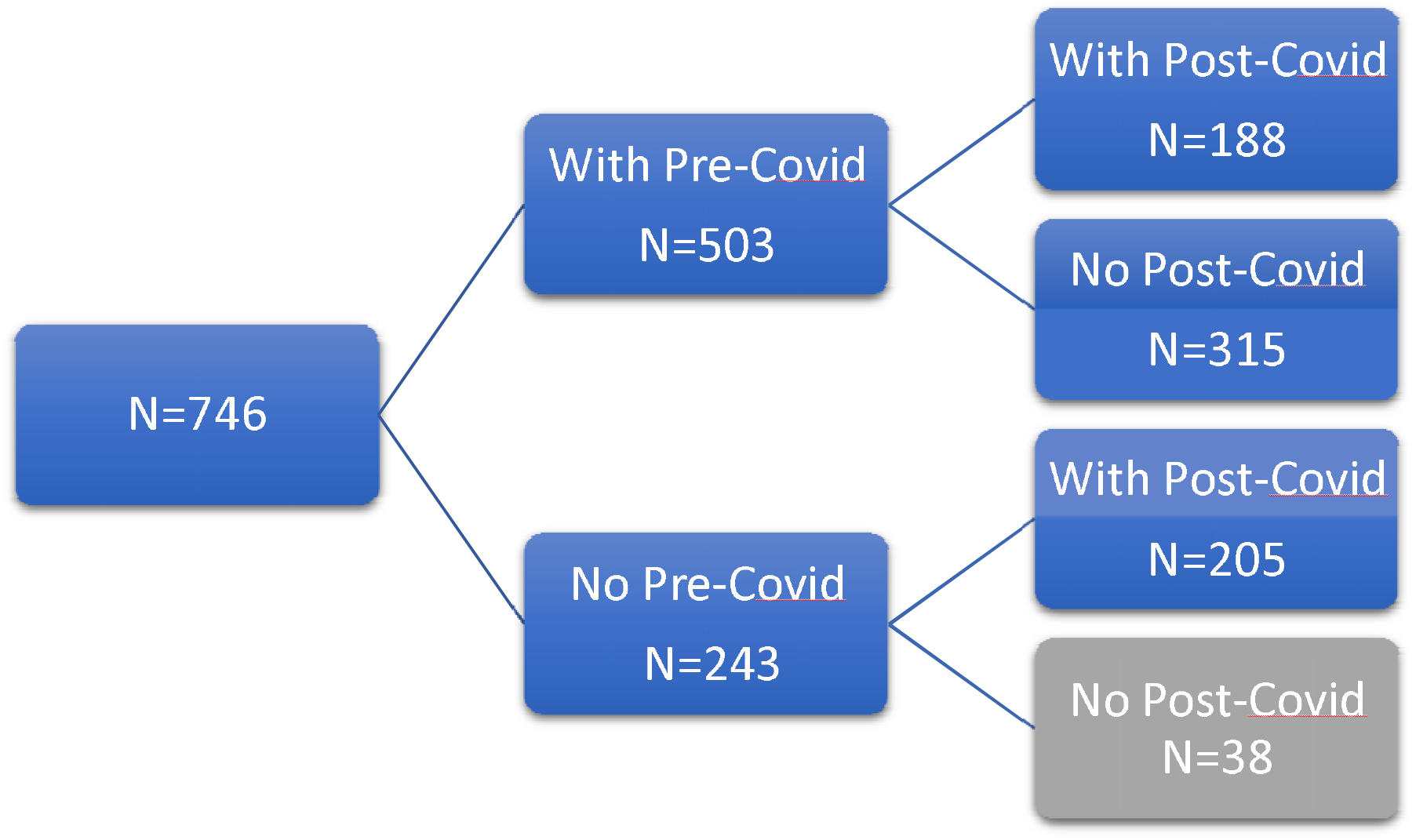
Sample flow chart of the total G2 sample of 746, which included all who had *either* pre or during COVID-19 data and excluded those with no pre or during COVID-19 data (n = 38).

**Figure S2.**
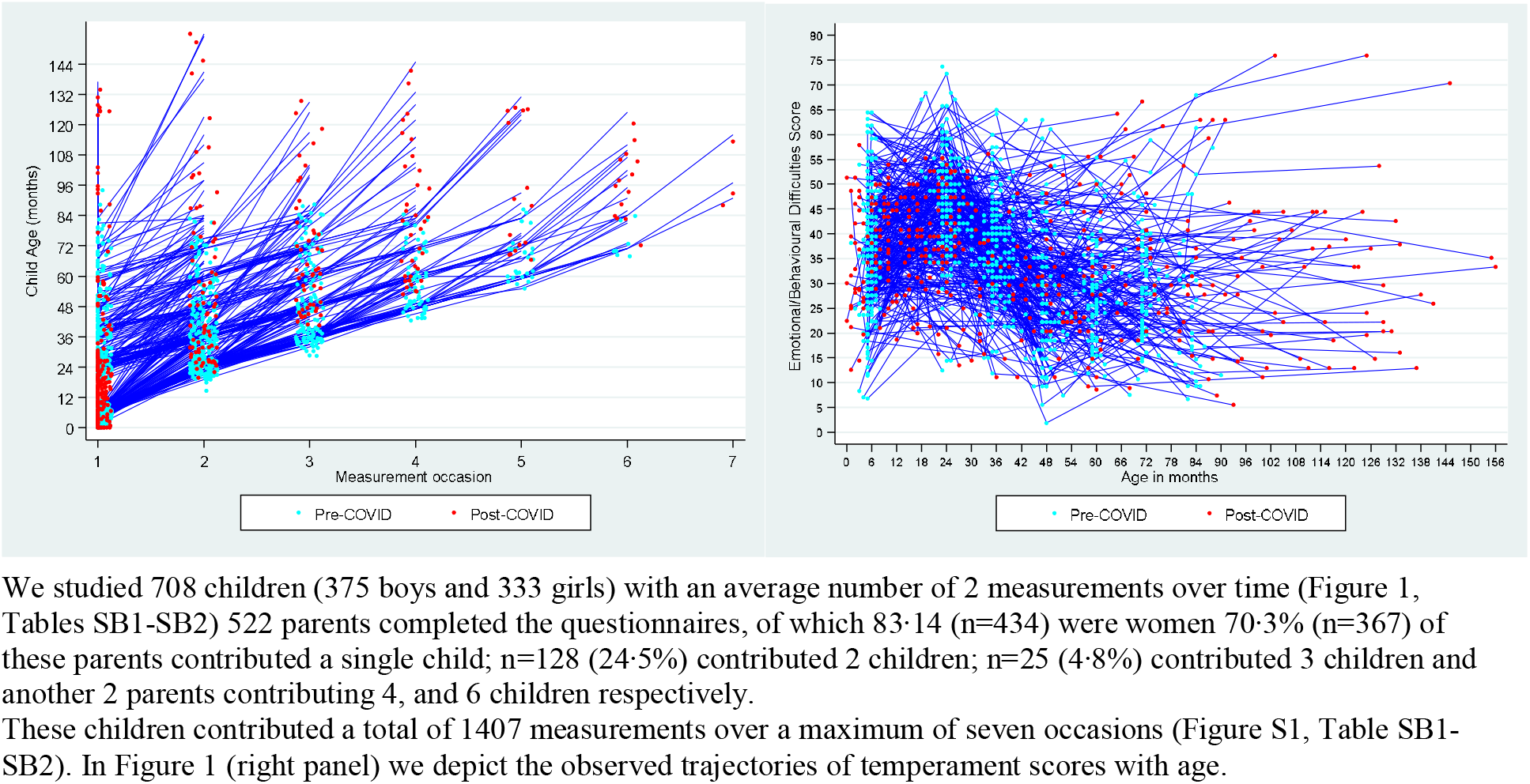
Description of follow-up observations over time and observed trajectories of children’s emotional and behavioural difficulties.

**Figure S3.**
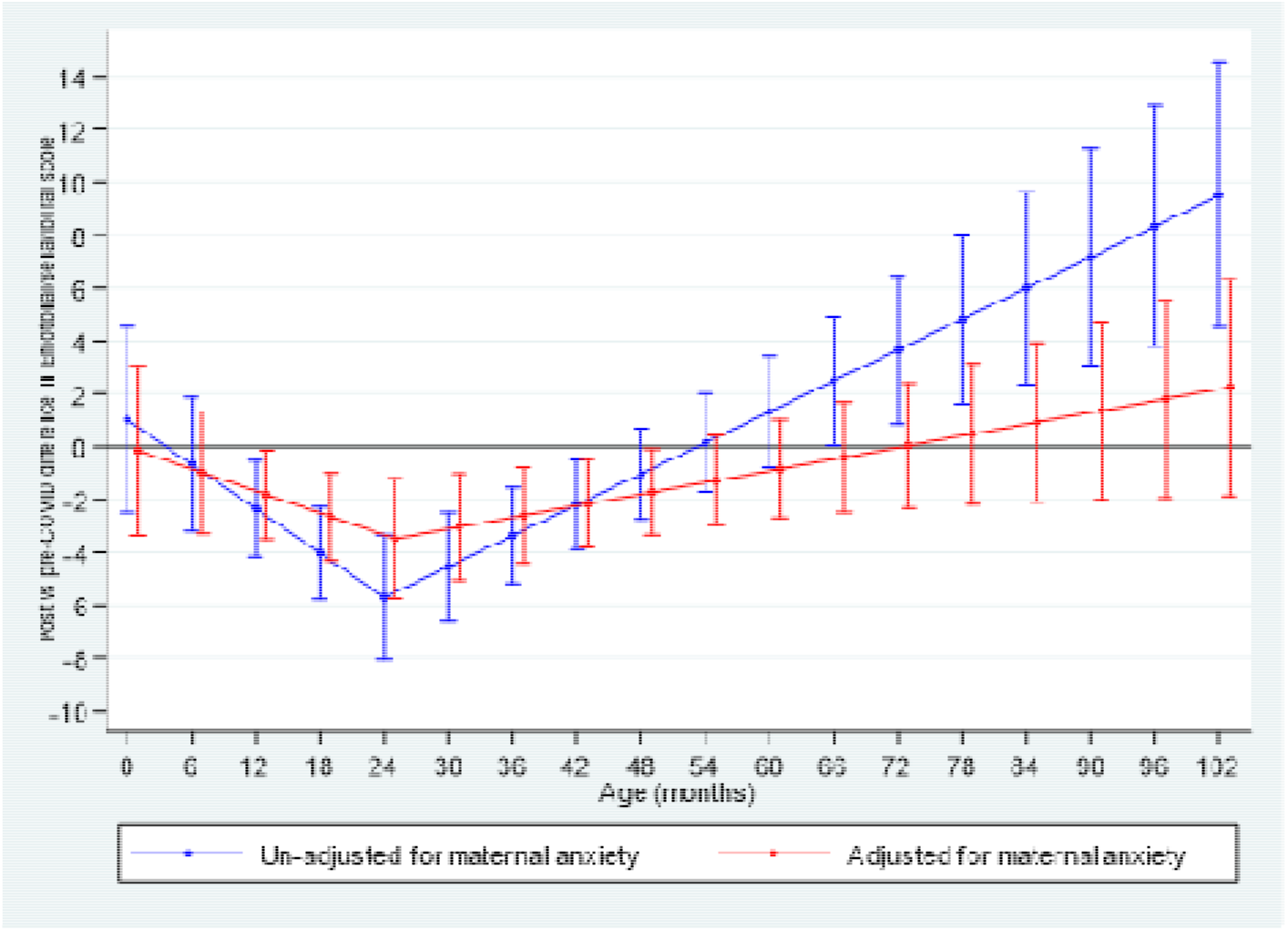
Pandemic differences in children’s emotional and behavioural difficulties adjusted and unadjusted for parental anxiety.

**Figure S4.**
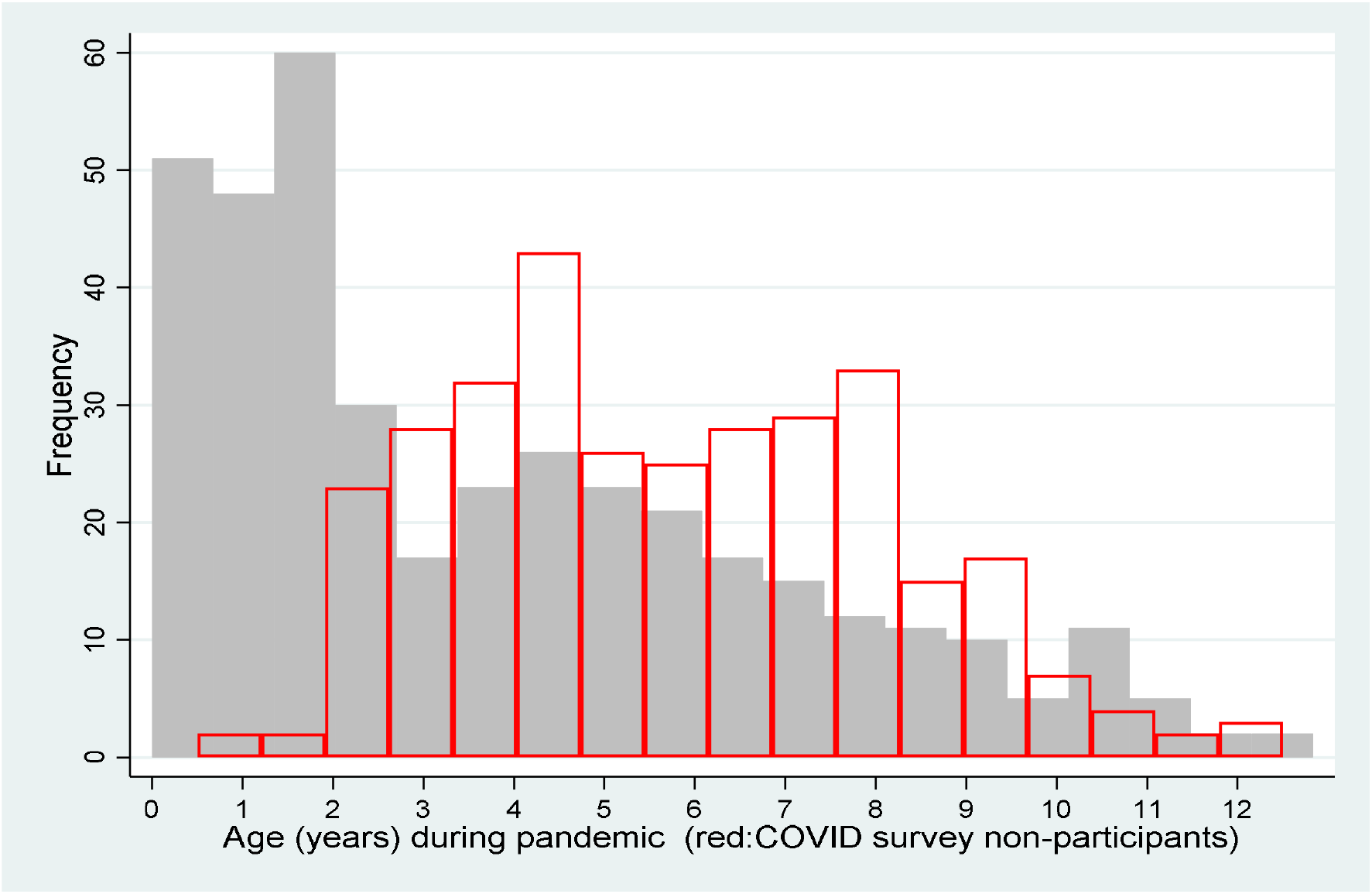
Distribution of child age (years) during the pandemic. Greyed are the children who contribute observation during the pandemic n=393 and red is those who are in G2 but didn’t return the COVID survey n=315. This age also represents the follow-up time for those participating and contribute to our results. Participants in the COVID survey are also children with less follow-up time as there were not old enough to contribute but there is still considerable overlap. This is taken into account when examining rates of missing visits.

**Figure S5.**
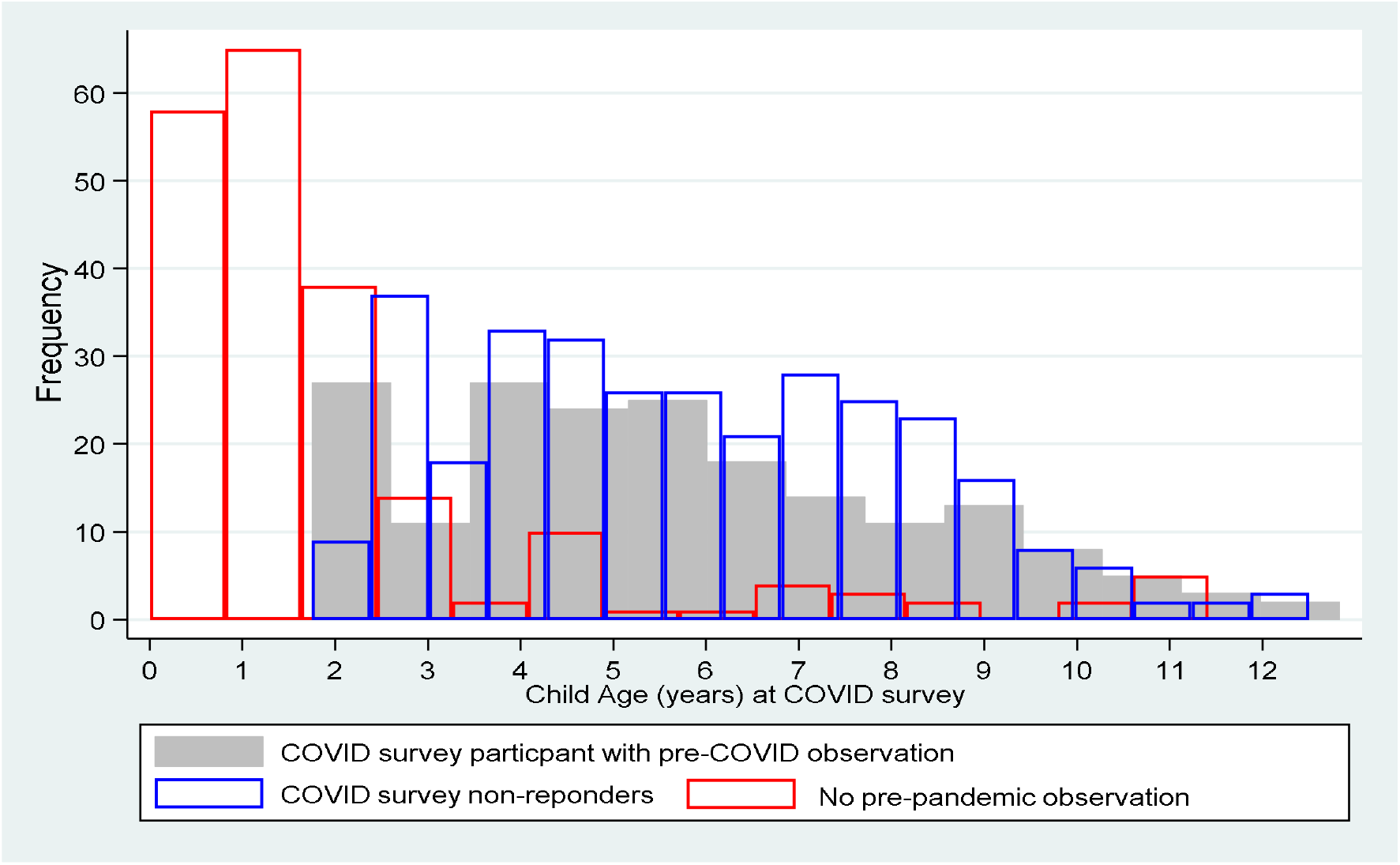
Distribution of child age (years) during the pandemic. Greyed are the children who contribute observation during the pandemic and pre-pandemic n=188, blue is those who are in G2 but didn’t return the COVID survey n=315 and red is those who returned the pandemic survey but do not have pre-pandemic data.

**Figure S6.**
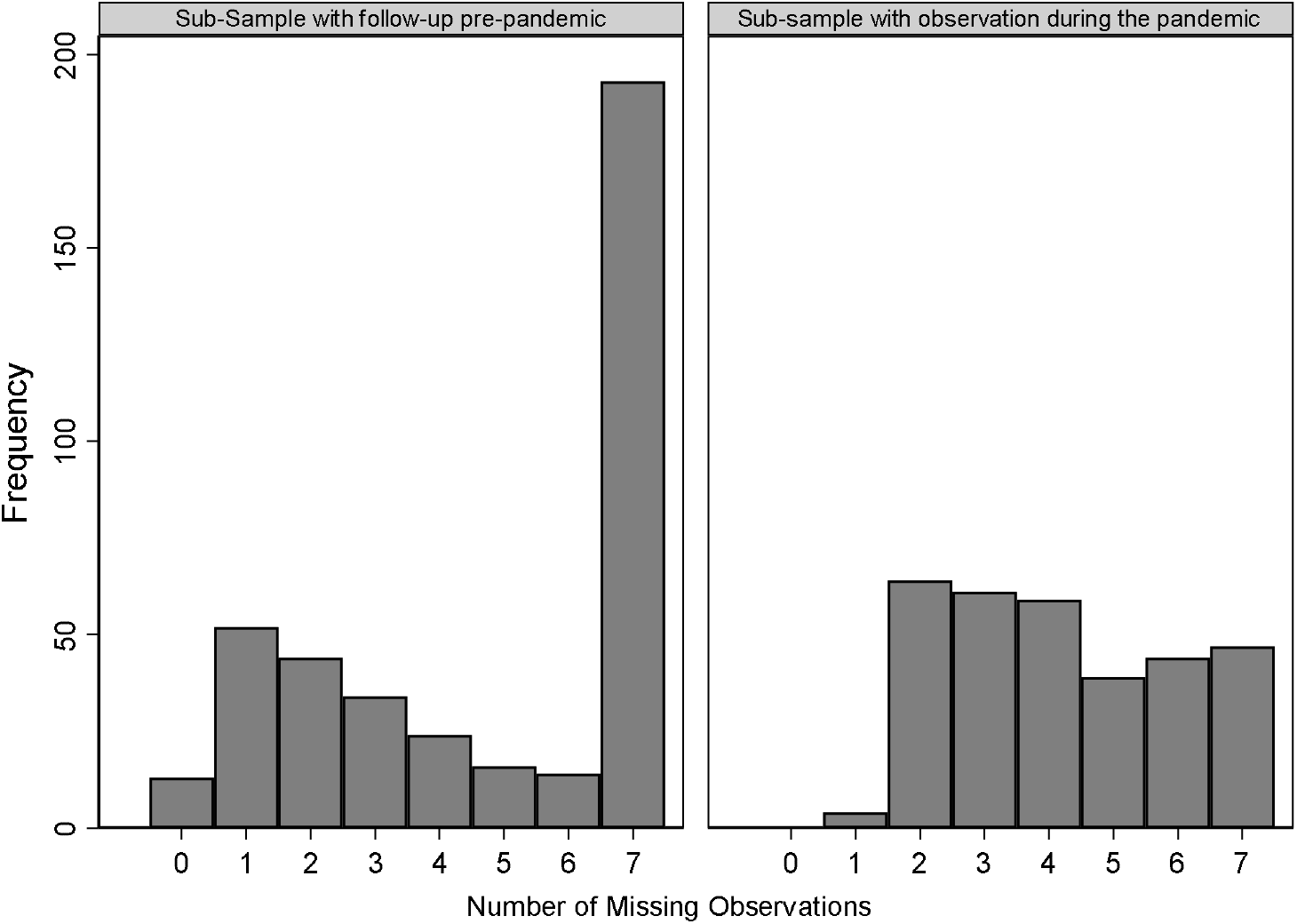
A description of the number of missing observations between those with and without observation pre-pandemic. The number of missing visits are calculated on the basis of visits a child could have had based on their age at the time of the G2 survey. For example, a child participating in the G2 survey who was 2 years old at the time of the survey could have had a maximum of two visits (could have participated in the pre-pandemic waves at 6 months) and no more than one missing observation.

**Figure S7.**
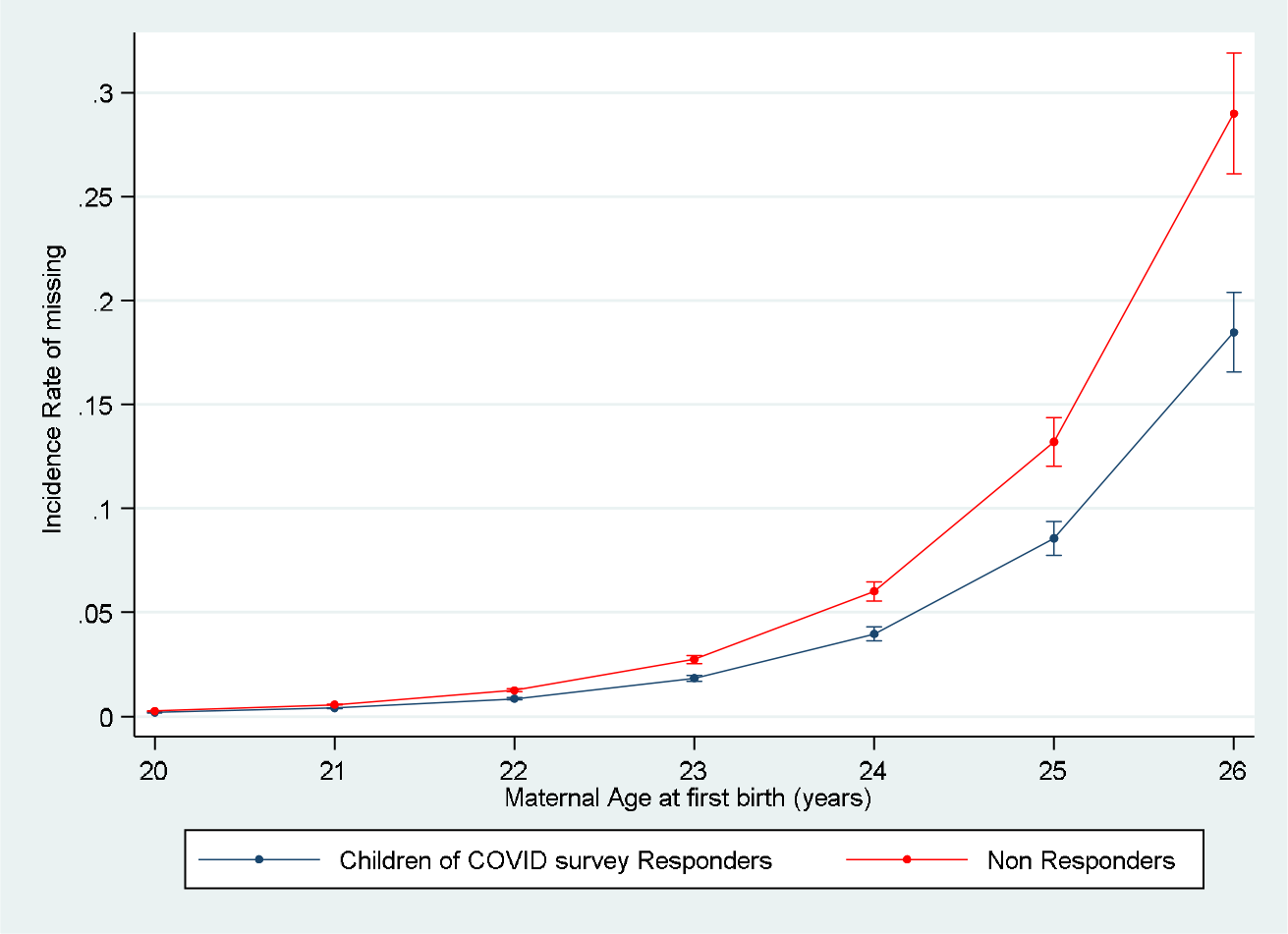
A depiction of how missing rates are influenced by maternal age between responders and non-responders to the COVID survey, where it can be seen how missing rates almost double with every year of maternal age at birth of first child, along with the higher missing rates for non-responders. We found no evidence of a difference in the effect of maternal age on missing rates between responders and non-responders. The IRR for missing rates according to a year increase in maternal age is 2·15 (95% CI= [2.13 to 2.18]) among the children of survey responders vs 2·19 (95% CI= [2·17 to 2·22]) in the children of survey non-responders.

